# Associations Between Depression, Anxiety, and Multiple Disease Outcomes: A Phenotype x Phenome-Wide Association Study in Electronic Health Records

**DOI:** 10.1101/2022.06.03.22275969

**Authors:** Y. Nina Gao, Brandon Coombes, Euijung Ryu, Vanessa Pazdernik, Gregory Jenkins, Richard Pendegraft, Joanna Biernacka, Mark Olfson

**Affiliations:** Department of Psychiatry, Columbia University and New York State Psychiatric Institute, New York, USA; Department of Quantitative Health Sciences, Mayo Clinic, Rochester, MN, USA; Department of Psychiatry and Psychology, Mayo Clinic, Rochester, MN, USA

**Author notes:** These authors contributed equally to this work (Y. Nina Gao, Brandon Coombes). These authors contributed equally to this work (Joanna Biernacka, Mark Olfson).

## Abstract

Anxiety and depression are frequently comorbid yet phenotypically distinct. This study identifies differences in the clinically observable phenome across a wide variety of physical and mental disorders comparing patients with diagnoses of isolated depression, isolated anxiety, or both. Using electronic health records for 14,994 participants with depression and/or anxiety in the Mayo Clinic Biobank, a phenotype x phenome-wide association study (Phe^2^WAS) was performed to test for differences between these groups. Additional Phe^2^WAS were performed restricting analyses to diagnoses occurring before or after a diagnosis of depression or anxiety to determine the temporal sequencing of diagnoses. Compared to isolated anxiety, isolated depression was more strongly associated with obesity (OR=1.61; p=3e-22), sleep apnea (OR=1.71; p=1e-22), type II diabetes (OR=1.74; p=9e-18), and neurological disorders (OR=1.54; p=6e-10); these differences were most pronounced for medical diagnoses made after first depression or anxiety diagnosis. In relation to isolated depression, isolated anxiety was more strongly related to palpitations (OR=1.91; p=2e-25), skin neoplasms (OR=1.61; p=2e-17), cardiac dysrhythmias (OR=1.45; p=2e-12), and degenerative skin conditions (OR=1.41; p=1e-10); these differences were most pronounced for medical diagnoses made before a first depression or anxiety diagnosis. Patients with the comorbid depression and anxiety had stronger associations with other mental health disorders, substance use disorders, sleep problems, and digestive problems relative to those with isolated depression or anxiety. While depression and anxiety are closely related, this study suggests that improving phenotypic characterization within the broad categories of depression and anxiety could improve clinical care and provide future directions for research.

## Introduction

Although anxiety and depression are typically differentiated in specialty mental health settings, these diagnoses are frequently comorbid [1]. Pharmacologically, the two disorders are treated similarly with agents chiefly active on the serotoninergic system. Genetic studies further suggest almost complete overlap between anxiety and depression [2, 3]. However, despite these similarities, and the consequent hypotheses of a shared etiology [4–6], we continue to have a limited understanding of why certain individuals develop anxiety disorders, others develop depressive disorders, and many develop both.

Heterogeneity, both between and within each of these disorders, contributes to the difficulty of characterizing them both genetically and neurobiologically. Under the Research Domain Criteria (RDoC) project, depression has been characterized as a disorder of impaired reward response, learning, and valuation [7, 8]. By contrast, anxiety has been conceptualized in RDoC as a dysfunction of threat detection [7, 8]. Clinically, however, there may be several overlapping domains. Anxiety symptoms have been incorporated into some diagnostic criteria for depression [9]. Other clinical symptoms such as psychomotor disturbances, appetite alterations, sleep patterns, and suicidal behaviors have also been incorporated into recent work attempting to identify genetically distinct depression subtypes with the hope of identifying neurobiological therapeutic targets [10].

Because depression and anxiety are both common mental disorders, they have been widely studied in medical settings [11–14]. Both disorders have been found to be associated with functional impairment and adverse outcomes across multiple settings ranging from increased disability and days off from work [14] to elevated risk for cardiovascular events and lower rates of cardiovascular recovery [12]. The distinctions between symptomatic phenotypes within and between these two disorders are also medically meaningful. Some evidence suggests depression and anxiety vary in the strength of their correlations with specific physical conditions. Compared to anxiety, for example, depression has a stronger correlation with obesity, while metabolic and inflammatory conditions are strongly correlated with hyperphagia, hypersomnia, and lethargy [11]. Similarly, skin diseases have been associated with panic severity, but not with depression severity [15]. As patients age, and physical health declines, new associations may further emerge between a patient’s mental health symptoms and physical conditions.

While factors such as the timing of onset have been found to differentiate depression subtypes [10, 16], it has not been frequently incorporated into exploratory investigations. This is partly due to the design of many prior studies, which have addressed specific hypotheses regarding single conditions or sought to describe lifetime risk [17]. However, temporal factors, such as the age at onset of illness or the timing of illness onset relative to the onset of comorbidity, can be inform understanding of clinical course. For example, anxiety disorders are much more likely to present at younger ages and to predate diagnosis of depressive disorders [18, 19]; however, new anxiety, particularly presenting in older populations, may be indicative of new cognitive impairment or dementia [16].

The relationship between mental and physical disorders is often further complicated by bidirectionality. Observed comorbidity rates of depression and anxiety exceed what would have been predictable by heritability suggesting common environmental stressors as well as common genetic preloading [10, 19]. Adverse health outcomes can also themselves act as stressors and contribute psychologically and physiologically to symptoms of depression and anxiety [20, 21]. With these considerations in mind, we conducted an exploratory analysis to identify differences in the clinically observable phenome, representing a wide variety of mental and physical health disorders, between patient with diagnoses of depression, anxiety, or both. Using the Mayo Clinic Biobank, we performed a phenotype x phenome-wide association study (Phe^2^WAS). PheWAS is an agnostic approach that originally involved testing for association between genetic variation or genetic risk across the clinical phenome observed in the electronic health record (EHR) as measured using diagnostic phecodes [17, 22]. Instead of using genetic data as the predictor, the Phe^2^WAS approach uses another phenotype as the predictor to identify phenotypic correlations across the EHR with the phenotype of interest or, if a binary phenotype, test for differences between two different groups of patients. We defined several clinical phenotypes within the broader categories of depression and anxiety (isolated depression, isolated anxiety, and comorbid depression and anxiety) and applied the Phe^2^WAS approach to test for differences between the clinical groupings across the EHR phenome (i.e. phecodes). We also examined the impact of temporality of other diagnoses vs. the anxiety/depression diagnoses on the Phe^2^WAS as a means of further differentiating the clinical groupings of isolated anxiety and isolated depression.

## Methods

Data for this study were derived from the EHR of participants in the Mayo Clinic Biobank [23]. All patients in the study gave informed consent for biobank research and the study was approved by the Mayo Clinic Institutional Review Board and the Mayo Clinic Biobank Access Committee.

The EHR includes clinical data as well as data on demographics, medications/prescriptions, laboratory values, billing codes from the International Classification of Diseases, 9^th^ and 10^th^ editions (ICD-9-CM and ICD-10-CM), and Current Procedural Terminology (CPT) codes [24]. We mapped ICD-9-CM and ICD-10-CM codes to phecodes (i.e. higher order group of diagnoses) as described and validated by the Phecode map 1.2b1 [25]. For the current study, we included all available EHR data from before April 6, 2020. Patients were defined as having a given phecode diagnosis if they had at least 2 instances of codes from that phecode in their EHR with the index date of each code defined as the first instance in the HER.

### Defining cases with MDD and ANX

Using structured EHR data, depression was defined as having at least two depression-related ICD codes on two different dates, using an initial list of codes mapped to phecodes for major depressive disorder (MDD) (phecodes 296.2 and 296.22), available from https://phewascatalog.org/phecodes [26, 27] with the addition of dysthymic disorder [ICD-9: 300.4; ICD-10: F34.1], depressive type psychosis [ICD-9: 298.0], and atypical depressive disorder [ICD-9: 296.82]. Anxiety was defined as having at least one anxiety-related code, using an initial list of codes mapped to phecodes for anxiety disorders (phecode: 300.1, 300.11, 300.12, 300.13), available from https://phewascatalog.org/phecodes [26, 27] with the addition of separation anxiety disorder (ICD-9: 309.21; ICD10: F93.0) and selective mutism (ICD-9: 313.23; ICD-10: F94.0) and removing phobic and social anxiety disorder in childhood (ICD-10: F93.1 and F93.2), hysteria (ICD-9: 300.1), anxiety disorder in conditions classified elsewhere (ICD-9: 293.84; ICD-10: F06.4) and overanxious disorder (ICD-9: 313). The complete lists of codes for defining MDD and anxiety are presented in Supplemental Tables S1 and S2. Patients with only 1 code for depression or anxiety were excluded from the analyses. Patients without codes for depression and anxiety were also excluded from the analyses.

### Phe^2^WAS

We first used logistic regression models to assess how the phenome of those with isolated depression differed from those with isolated anxiety (MDD-only vs. ANX-only) using a Phe^2^WAS approach. In this approach, the phenotype MDD-only vs. ANX-only was tested for association with each phecode in the EHR that was not rare (> 100 cases of the phecode in the analysis). To account for potential demographic confounders, we adjusted the analyses for current age, self-reported gender, race, and ethnicity. To account for potential confounding by length and depth of EHR, we also adjusted for length of record (defined as the length of time from a patient’s first ICD code (any ICD code, not limited to depression/anxiety) in their EHR to most recent ICD code), median age of record (median time from date of data pull to date of each ICD code in a patient’s record), and total number of ICD codes (defined as the total number of codes in a patient’s EHR while allowing for duplicated codes on different dates).

Physical and psychiatric symptoms can influence each other; for example, reduced mobility due to medical illness can potentiate subsequent depressive symptoms, or neurovegetative symptoms of depression can potentiate deconditioning and poorer health. Because temporality is important for characterizing illness phenotype and suggesting potential points of intervention, we repeated the analyses comparing MDD-only vs. ANX-only separately restricting to phecodes that occur before diagnosis of depression/anxiety or by restricting to phecodes that occur after diagnosis of depression/anxiety. In these analyses, we adjusted analyses for age at diagnosis of depression/anxiety instead of current age to account for the fact that we truncated the EHR. We also updated the length of record in these analyses to account for the truncated EHR.

Finally, we assessed whether the phenome of those with comorbid depression and anxiety (MDD+ANX) differed from those with only one disorder. To do this, MDD+ANX vs. only one disorder was tested for association with each phecode in the EHR with adjustments as described above for current age, self-reported gender, race, ethnicity, length of record, median age of record, and total number of ICD codes.

The significance threshold was set using a strict Bonferroni correction for the number of phecodes tested in each Phe^2^WAS (p < 0.05/1618 = 3e-5).

## Results

We identified 14994 patients from the Mayo Clinic Biobank with at least 2 codes for anxiety or depression. Of these, 5246 (35.0%) had isolated depression, 3213 (21.4%) had isolated anxiety, and 6535 (43.6%) had comorbid depression and anxiety. Proportions of isolated versus comorbid diagnosis differed between men and women: 46.9% of women in our sample were diagnosed with comorbid depression and anxiety compared to 36.1% of men. The sample was older (mean current age 66.4 years) and mostly non-Hispanic (95.0%), white (90.2%), and female (69.3%) with no large differences in demographics among the different diagnostic groups (Table 1). Of note, patients with isolated anxiety and isolated depression were diagnosed at an older age (mean age of 57.5 and 56.1 years old, respectively) than those with comorbid anxiety and depression (mean age of 52.3 and 50.2 years old, respectively). Furthermore, while those with isolated anxiety and depression had fewer total ICD codes in their record (mean=514 and 585, respectively) compared to patients with the comorbidity (mean=848 codes), all groups had similar length of record and median age of record indicating that those with the comorbidity could have higher rates of other comorbid conditions as well.

**Table 1.** Demographics and EHR description

| Variable | Level |  | All | MDD+ANX | MDD-only | ANX-only |
| --- | --- | --- | --- | --- | --- | --- |
|  |  |  | N = 14994 | N = 6535 | N = 5246 | N = 3213 |
| Gender | Male | N (%) | 4608 (30.7%) | 1664 (25.5%) | 1808 (34.5%) | 1136 (35.4%) |
|  | Female | N (%) | 10386 (69.3%) | 4871 (74.5%) | 3438 (65.5%) | 2077 (64.6%) |
| Race | White | N (%) | 13525 (90.2%) | 5824 (89.1%) | 4779 (91.1%) | 2922 (90.9%) |
|  | Black/African American | N (%) | 108 (0.7%) | 51 (0.8%) | 38 (0.7%) | 19 (0.6%) |
|  | Asian | N (%) | 96 (0.6%) | 34 (0.5%) | 29 (0.6%) | 33 (1.0%) |
|  | Native American/Alaskan Native | N (%) | 31 (0.2%) | 18 (0.3%) | 9 (0.2%) | 4 (0.1%) |
|  | Other | N (%) | 74 (0.5%) | 37 (0.6%) | 21 (0.4%) | 16 (0.5%) |
|  | Mixed | N (%) | 771 (5.1%) | 379 (5.8%) | 239 (4.6%) | 153 (4.8%) |
|  | Unknown/Missing | N (%) | 389 (2.6%) | 192 (2.9%) | 131 (2.5%) | 66 (2.1%) |
| Ethnicity | Non-Hispanic | N (%) | 14305 (95%) | 6205 (95%) | 5022 (96%) | 3078 (96%) |
|  | Hispanic | N (%) | 233 (1.6%) | 101 (1.6%) | 86 (1.6%) | 46 (1.4%) |
|  | Unknown | N (%) | 456 (3.0%) | 229 (3.5%) | 138 (2.6%) | 89 (2.8%) |
| Age | Current age (yrs old) | mean (SD) | 66.4 (16.3) | 63.9 (16.7) | 69.0 (15.5) | 67.1 (16.0) |
|  | Age at 1st Dx of ANX (yrs old) | mean (SD) | 54.0 (17.5) | 52.3 (17.5) | n/a | 57.5 (16.8) |
|  | Age at 1st Dx of MDD (yrs old) | mean (SD) | 52.8 (17.6) | 50.2 (17.6) | 56.1 (17.0) | n/a |
| Record | Median age of record (yrs) | mean (SD) | 8.24 (3.7) | 8.0 (3.65) | 8.9 (3.76) | 7.64 (3.54) |
|  | Length of record (yrs) | mean (SD) | 21.5 (10.7) | 22.4 (10.4) | 21.4 (11.0) | 19.7 (10.6) |
|  | Number of Dx codes in EHR | mean (SD) | 684 (819) | 848 (1000) | 586 (631) | 514 (591) |

Legend: MDD = depression; ANX = anxiety; current age = age at date of data extraction; Median age of record = median age of all ICD codes in a patient's record

### Phe^2^WAS of isolated depression versus isolated anxiety

In relation to patients with isolated depression, patients with isolated anxiety had notably higher frequency of obesity (OR=1.61; p=3e-22), sleep apnea (OR=1.71; p=1e-22), type II diabetes (OR=1.74; p=9e-18), schizophrenia (OR=16.5; p=2e-16), and neurological disorders (OR=1.54; p=6e-10). In our models to assess the temporality of these findings, we found that the higher comorbidity frequencies of obesity, schizophrenia, and neurological disorders in those with isolated depression were only significantly higher if we restricted to phecodes occurring after diagnosis of depression or anxiety (Supplemental Table 3), which may imply these differences between isolated depression and isolated anxiety were most pronounced for medical diagnoses made after the first diagnosis of depression or anxiety. Differences in the groups found with sleep apnea and type II diabetes persisted regardless of truncation.

We also found that rates of many conditions including palpitations (OR=1.91; p=2e-25), neoplasms of the skin (OR=1.61; p=2e-17), cardiac dysrhythmias (OR=1.45; p=2e-12), degenerative skin conditions (OR=1.41; p=1e-10), and dyspepsia (OR=1.51; p-8e-8) were higher in patients with isolated anxiety than those with isolated depression. After truncating the phecodes to either before or after diagnosis, we found that the higher comorbidity rates of neoplasms of the skin, cardiac dysrhythmias, and degenerative skin conditions were only significantly higher in those with isolated anxiety if we restricted to phecodes occurring before diagnosis of anxiety or depression (Supplemental Table 3). This indicates that these differences were most pronounced for medical diagnoses made before the first diagnosis of depression or anxiety. Differences between the groups in rates of palpitations and dyspepsia persisted regardless of truncation.

### Phe^2^WAS of comorbid depression and anxiety versus those with only one disorder

In comparing the lifetime diagnostic phecodes of patients with comorbid depression and anxiety to those with isolated depression or anxiety (Figure 2), patients with the comorbidity were more likely to have other mental health disorders, substance use disorders, sleep problems, digestive issues, and symptoms. In particular, among many codes, rates of insomnia (OR=1.70; p=1e-39), suicidal ideation or attempt (OR=4.87; p=5e-38), post-traumatic stress disorder (OR=3.5; p=8e-32), substance use disorders (OR=2.15; p=4e-23), alcohol use disorder (OR=2.02; p=2e-22), chronic pain syndrome (OR=1.72; p=2e-16), GERD (OR=1.32; p=2e-13), irritable bowel syndrome (OR=1.42; p=2e-9) and myalgia and myositis (OR=1.4; p=2e-16) were higher in those with the comorbidity than those with isolated anxiety or depression.

**Figure 1.**
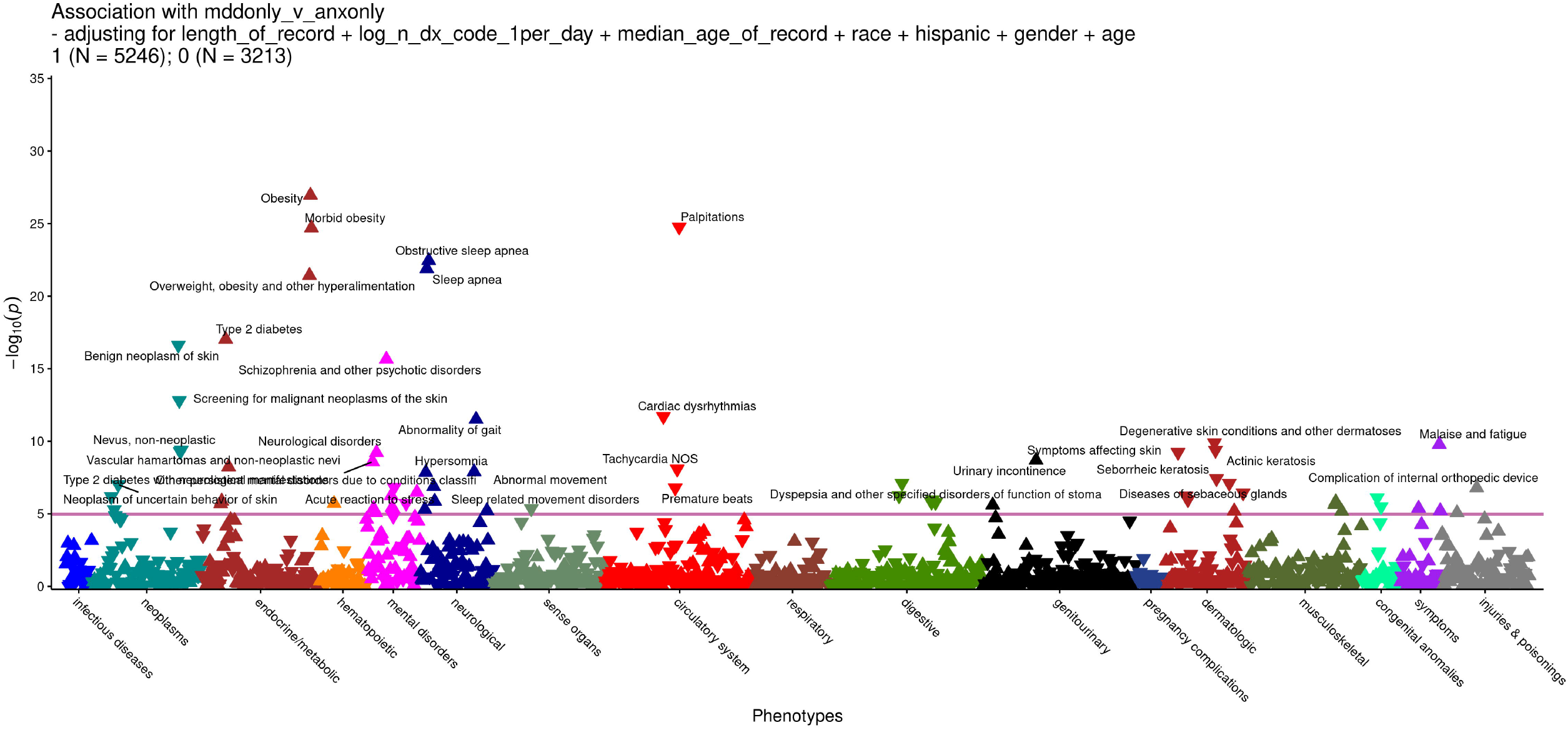
Manhattan plot comparing the phenome of those with isolated depression to those with isolated anxiety. Triangles with single point upward indicate phecodes more commonly observed in patients with isolated depression than in patients with isolated anxiety. Triangles with single point downwards indicate phecodes more common in patients with isolated anxiety than in those with isolated depression. Associations are adjusted for current age, self-reported gender, race, and ethnicity, length of EHR record, median age of all ICD codes in a patient’s record, and total number of ICD codes. Participants with comorbid depression and anxiety, and those with neither condition, are excluded from this analysis.

**Figure 2.**
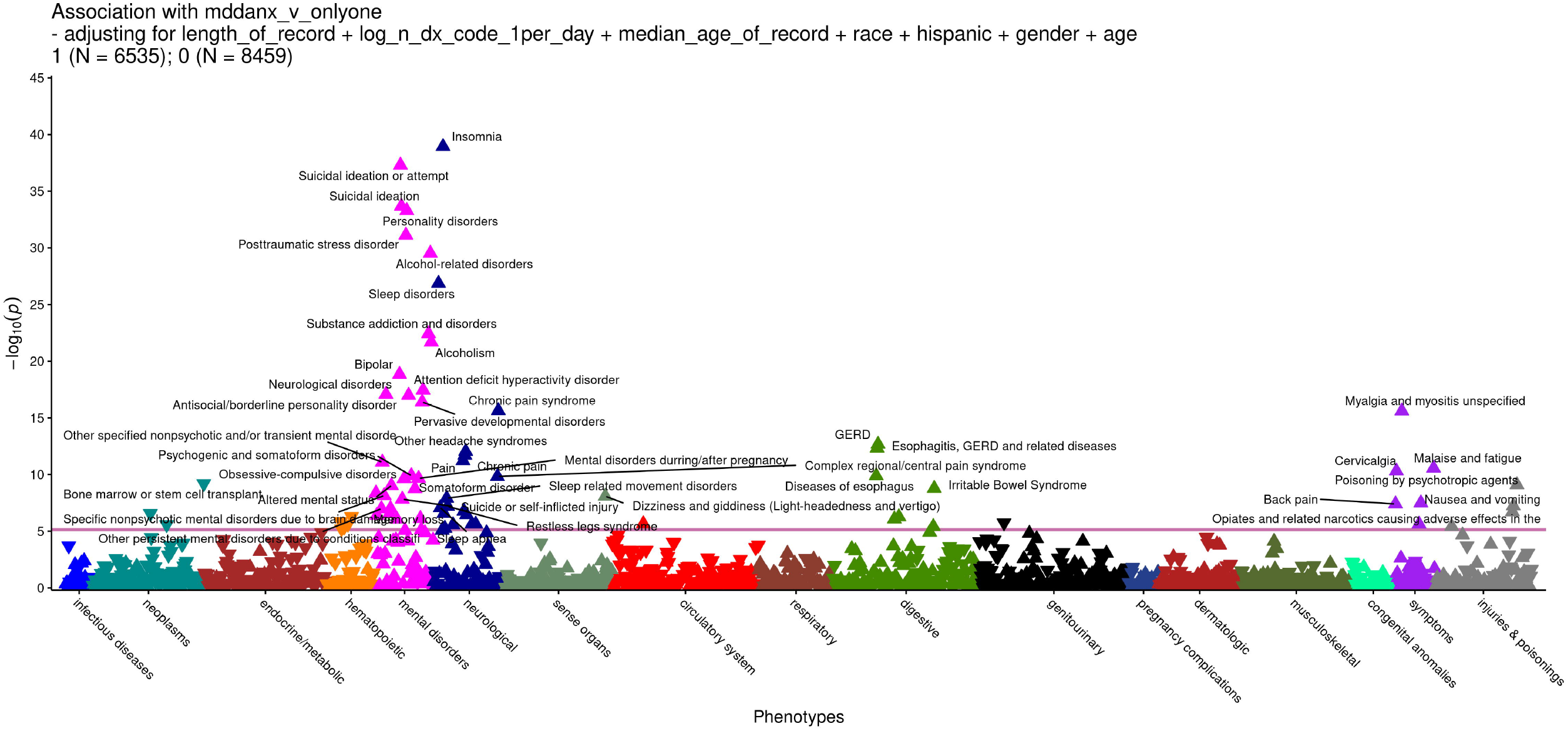
Manhattan plot comparing the lifetime phenome of those with comorbid depression and anxiety to those with isolated depression or anxiety. Triangles with single point upward indicate phecodes more commonly associated with comorbid depression and anxiety as compared to either isolated depression or isolated anxiety. Triangles with single point downwards indicate phecodes more commonly occurring with either isolated depression or isolated anxiety than with comorbid depression and anxiety. Analyses are adjusted for current age, self-reported gender, race, and ethnicity, length of EHR record, median age of record, and total number of ICD codes.

## Discussion

In contrast to prior studies, we defined phenotypes of isolated depression and isolated anxiety in addition to those with comorbid depression and anxiety [17, 18]. Compared to patients with isolated anxiety, patients with isolated depression had a higher frequency of metabolic abnormalities such as obesity and type II diabetes, as well as sleep apnea and hypersomnia. Differences between rates of obesity, schizophrenia, and neurological disorders were especially pronounced for diagnoses that were made *after* the initial depression or anxiety diagnosis. By contrast, the isolated anxiety group had significant overlaps with dermatologic diagnoses such as benign neoplasms and degenerative skin disorders, as well as cardiac symptoms such as palpitations and dysrhythmia; these differences were especially pronounced for medical diagnoses made *before* the first diagnosis of depression or anxiety.

These findings have several implications for our understanding of depression and anxiety and their relationships with other organ systems. First, consistent with prior research, we found a subset of patients with both depressive and gastrointestinal disorders [28–31]. The autonomic nervous system and serotonergic systems are implicated in gastrointestinal functions: gastric smooth muscle cells express 5-HT1 and 5-HT2 receptors where serotonin induces contractions in the gastric fundus and activity at 5-HT3 receptors caused gastric relaxation and accelerated intestinal transit [32, 33]. The common gastrointestinal side effects associated with SSRI antidepressants are illustrative of this relationship, however, so too are reports that ondansetron, a 5-HT3 receptor antagonist, improves depressive symptoms [32, 34]. These, at times, contradictory claims may suggest that multiple phenotypes occur within the subpopulation of patients presenting with co-occurring depressive and gastrointestinal disorders, and that perhaps even within individuals, relationships between a given set of symptoms and single neurotransmitter activity may be nonlinear. Our findings suggest that despite the potentially extensive relationship between central nervous and gastrointestinal systems, there is also some specificity and that patients with concurrent depression and anxiety appear to be at particular risk, which may narrow the set of potential genetic or molecular mechanisms involved.

Second, a less explored relationship between anxiety and skin disorders also implicates autonomic nervous system and serotonergic responses [35–37]. 5-HT2A is found in the upper epidermis of skin and 5-HT3 is expressed in the basal epidermal layer [38, 39]. Topical doxepin, used orally as an antidepressant, has been used to treat cold urticaria [40]. Our study describes a phenotype of concurrent isolated anxiety and skin disorders. Although it is possible that temperamental anxiety might provoke treatment seeking, it is significant that we observed a specific association with dermatologic diagnoses over other previously reported associations between anxiety and conditions that might mimic anxiety symptoms such as asthma [41, 42]. Similarly, although a variety of conditions such as a positive screening mammogram or prostate-specific antigen, might provoke reactive anxiety, the association with anxiety and skin conditions appears especially strong. Although inflammation has been generally implicated in the association of skin disorders with psychiatric symptoms, inflammation is systemic [5]. Thus, we add to the literature on phenotypic specificity.

Third, we further describe a previously characterized phenotype involving isolated depression associated with obesity and sleep apnea [11]. Compared to the comorbid depression group, the isolated depression group exhibited lower concurrent psychiatric risk. However, isolated depression diagnosis preceded obesity and related chronic health sequelae [12, 17]. Although we cannot causally attribute obesity to isolated depression, the diagnosis of depression may present a clinical opportunity for intervention prior to the development of other serious health conditions.

Fourth, associations of isolated depression with schizophrenia or neurological disorders were particularly pronounced relative to isolated anxiety in the period after first depression diagnosis. Prodromal depression has been described in schizophrenia and neurodegenerative disorders such as Parkinson’s disease, Alzheimer’s disease, and vascular dementia [43–45]. Given the literature on the emergence of depression following neurological insults such as stroke or traumatic brain injury [46, 47], the strength of this temporal relationship between depression and neurological illness may be surprising, however, the population prevalence of depressive symptoms is high relative to that of specific neurological diagnoses or schizophrenia [48–50]. Possible initial misattribution of depressive symptoms to a primary mood disorder can also impact clinical care for the patients diagnosed with these disorders.

The results from these Phe^2^WAS analyses indicate that comorbid diagnoses of depression and anxiety are highly associated with many other psychiatric conditions including suicidal behaviors, substance use disorders, and personality disorders, even compared to those with isolated depression or anxiety, which is consistent with prior literature [17, 51, 52]. Among patients with depression diagnoses, those with comorbid depression and anxiety still outnumbered isolated depression, thus it is not surprising that aggregated studies conducted with samples of patients with depression are dominated by this subset. Depression as compared to anxiety was associated with gastroesophageal reflux, esophagitis, and gastritis, which parallels previous studies of depression compared to controls [17], though the rates of these conditions were even higher among those with comorbid depression and anxiety.

Most patients with depression or anxiety presented with either isolated depression or isolated anxiety. In previous studies, the reported rate of comorbid anxiety with depression has been as high as 85% of patients with depression and 90% of patients with anxiety [1]. However, our estimates are not out of line with estimates from other biobank samples [10]. Because the average median record length was 8.2 years, one explanation for this discrepancy is that, relative to surveys, EHRs are less sensitive at detecting lifetime psychiatric diagnoses. However, the phenotypic profiles of patients with isolated depression and isolated anxiety differed significantly from those with both depression and anxiety suggesting that these phenotypes correspond to distinct risk groups [53]. This aligns with a recent genetic study of depression, anxiety and their comorbidity, which showed independent depression and anxiety polygenic contributions to the comorbidity. Relative to prior analyses of epidemiological samples of non-elderly individuals, aged 15 to 54 years [18, 19], our sample was also older, with a median age of 66.4 years old. Some studies of older individuals have described both decreasing rates of depression and anxiety with increasing age and decreasing rates of comorbidity [16, 54]. Presenting constellations of depression and anxiety symptoms change with increasing age, reflecting cohort factors such as healthy survivor bias, social factors such as reactions to a loss of physical independence or social connections, and biological factors such as psychiatric symptoms of age-related cognitive decline [16, 54–56]. Our study results may reflect the evolving heterogeneity of depression and anxiety across the adult lifespan.

## Limitations

This study has several limitations. First, our analysis was based on data from the Mayo Clinic Biobank, which represents a selective population of older adults of primarily European descent with higher-than-average levels of education [57]. Second, this analysis, which examines phenotypes from the EHR, is limited by length of record, selection into treatment, and other recording variables. This limits the examination of depression/anxiety onset, which often occurs at ages younger than those captured in our EHR. Third, although our final sample included 14,994 observations, we were restricted by sample size to investigating associations between relatively prevalent conditions. As such, we cannot comment on granular relationships between specific diagnoses. Fourth, our analysis was limited to phenotypes. Although we attempted to elucidate temporal clinical presentations of patients, we cannot use these findings to draw direct causal inferences and therefore view the results as exploratory.

## Conclusions

PheWAS approaches, which invert the idea of genome-wide association studies, have been successful in demonstrating that specific genetic variations are associated with multiple traits [22]. Here, we leveraged this approach to instead examine associations with clinical groups of patients with depression and anxiety by using a phenotype, rather than a genotype, as a predictor of multiple clinical conditions derived from the EHR (Phe^2^WAS). The current results provide several interesting perspectives on the clinical landscape of depression and anxiety. Patients with comorbid depression and anxiety are a well-characterized psychiatric patient population. Due to their level of psychiatric risk, this group is also the most likely to present for mental health treatment at a relatively early age. By contrast, patients with isolated depression or anxiety may differentially present to primary care or a medical specialist, such as a cardiologist or a dermatologist. These differences have the potential to affect recruitment into clinical research and thus affect the generalizability of the results [58]. Learning from broad descriptive cohorts across service settings could help us better understand clinical heterogeneity, improve generalizability of diagnostic characterizations, and offer better treatments to patients with these common psychiatric disorders.

## Data Availability

All data produced in the present study are available upon reasonable request to the authors.

## Acknowledgements

This work was supported by a grant from the NIH (R01 MH121924). YNG was supported in part by a Moynihan Clinical Research Fellowship from the Leon Levy Foundation and Award Number R25MH086466 from the National Institute of Mental Health.

---

**Supplemental Table 1.** Depression definition

| Diagnosis | ICD9/10 | ICD | Phecode | Description |
| --- | --- | --- | --- | --- |
| MDD | 296.2 | ICD9 | 296.22 | Major depressive disorder, single episode |
| MDD | 296.2 | ICD9 | 296.22 | Major depressive disorder, single episode, unspecified degree |
| MDD | 296.21 | ICD9 | 296.2 | Major depressive disorder, single episode, mild degree |
| MDD | 296.22 | ICD9 | 296.22 | Major depressive disorder, single episode, moderate degree |
| MDD | 296.23 | ICD9 | 296.22 | Major depressive disorder, single episode, severe degree, without mention of psychotic behavior |
| MDD | 296.24 | ICD9 | 296.22 | Major depressive disorder, single episode, severe degree, specified as with psychotic behavior |
| MDD | 296.25 | ICD9 | 296.22 | Major depressive disorder, single episode, in partial or unspecified remission |
| MDD | 296.26 | ICD9 | 296.22 | Major depressive disorder, single episode in full remission |
| MDD | 296.3 | ICD9 | 296.22 | Major depressive disorder, recurrent episode |
| MDD | 296.3 | ICD9 | 296.22 | Major depressive disorder, recurrent episode, unspecified degree |
| MDD | 296.31 | ICD9 | 296.2 | Major depressive disorder, recurrent episode, mild degree |
| MDD | 296.32 | ICD9 | 296.22 | Major depressive disorder, recurrent episode, moderate degree |
| MDD | 296.33 | ICD9 | 296.22 | Major depressive disorder, recurrent episode, severe degree, without mention of psychotic behavior |
| MDD | 296.34 | ICD9 | 296.22 | Major depressive disorder, recurrent episode, severe degree, specified as with psychotic behavior |
| MDD | 296.35 | ICD9 | 296.22 | Major depressive disorder, recurrent episode, in partial or unspecified remission |
| MDD | 296.36 | ICD9 | 296.22 | Major depressive disorder, recurrent episode, in full remission |
| MDD | 311 | ICD9 | 296.2 | Depressive disorder NEC |
| MDD | 300.4 | ICD9 | 300.4 | Dysthymic disorder |
| MDD | 298.0 | ICD9 | 295.3 | Depressive type psychosis |
| MDD | 296.82 | ICD9 | 296.1 | Atypical depressive disorder |
| MDD | F32 | ICD10 | 296.22 | Major depressive disorder, single episode |
| MDD | F32.0 | ICD10 | 296.22 | Major depressive disorder, single episode, mild |
| MDD | F32.1 | ICD10 | 296.22 | Major depressive disorder, single episode, moderate |
| MDD | F32.2 | ICD10 | 296.22 | Major depressive disorder, single episode, severe without psychotic features |
| MDD | F32.3 | ICD10 | 296.22 | Major depressive disorder, single episode, severe with psychotic features |
| MDD | F32.4 | ICD10 | 296.22 | Major depressive disorder, single episode, in partial remission |
| MDD | F32.5 | ICD10 | 296.22 | Major depressive disorder, single episode, in full remission |
| MDD | F32.8 | ICD10 | 296.22 | Other depressive episodes |
| MDD | F32.89 | ICD10 | 296.22 | Single episode of 'masked' depression NOS |
| MDD | F32.9 | ICD10 | 296.22 | Major depressive disorder, single episode, unspecified |
| MDD | F33 | ICD10 | 296.22 | Major depressive disorder, recurrent |
| MDD | F33.0 | ICD10 | 296.22 | Major depressive disorder, recurrent, mild |
| MDD | F33.1 | ICD10 | 296.22 | Major depressive disorder, recurrent, moderate |
| MDD | F33.2 | ICD10 | 296.22 | Major depressive disorder, recurrent severe without psychotic features |
| MDD | F33.3 | ICD10 | 296.22 | Major depressive disorder, recurrent, severe with psychotic symptoms |
| MDD | F33.4 | ICD10 | 296.22 | Major depressive disorder, recurrent, in remission |
| MDD | F33.40 | ICD10 | 296.22 | Major depressive disorder, recurrent, in remission, unspecified |
| MDD | F33.41 | ICD10 | 296.22 | Major depressive disorder, recurrent, in partial remission |
| MDD | F33.42 | ICD10 | 296.22 | Major depressive disorder, recurrent, in full remission |
| MDD | F33.8 | ICD10 | 296.22 | Other recurrent depressive disorders |
| MDD | F33.9 | ICD10 | 296.22 | Major depressive disorder, recurrent, unspecified |
| MDD | F34.1 | ICD10 | 300.4 | Dysthymic disorder |

**Supplemental Table 2:** Anxiety definition

| **Diagnosis** | **ICD9/10** | **ICD** | **PheCode** | **ICD9 String** |
| --- | --- | --- | --- | --- |
| ANX | 300 | ICD9 | 300.1 | Anxiety states |
| ANX | 300 | ICD9 | 300.1 | Anxiety state unspecified |
| ANX | 300.01 | ICD9 | 300.12 | Panic disorder without agoraphobia |
| ANX | 300.02 | ICD9 | 300.11 | Generalized anxiety disorder |
| ANX | 300.09 | ICD9 | 300.1 | Other anxiety states |
| ANX | 300.2 | ICD9 | 300.13 | Phobia unspecified |
| ANX | 300.21 | ICD9 | 300.12 | Agoraphobia with panic disorder |
| ANX | 300.22 | ICD9 | 300.12 | Agoraphobia without mention of panic attacks |
| ANX | 300.23 | ICD9 | 300.12 | Social phobia |
| ANX | 300.29 | ICD9 | 300.13 | Other isolated or specific phobias |
| ANX | 309.21 | ICD9 | 313 | Separation anxiety disorder |
| ANX | 313.23 | ICD9 | 313 | Selective mutism specific to childhood and adolescence |
| ANX | F40 | ICD10 | 300.13 | Phobic anxiety disorders |
| ANX | F40.0 | ICD10 | 300.12 | Agoraphobia |
| ANX | F40.1 | ICD10 | 300.12 | Social phobias |
| ANX | F40.2 | ICD10 | 300.13 | Specific (isolated) phobias |
| ANX | F40.8 | ICD10 | 300.13 | Other phobic anxiety disorders |
| ANX | F40.9 | ICD10 | 300.13 | Phobic anxiety disorder, unspecified |
| ANX | F41.0 | ICD10 | 300.12 | Panic disorder [episodic paroxysmal anxiety] |
| ANX | F41.1 | ICD10 | 300.11 | Generalized anxiety disorder |
| ANX | F41.2 | ICD10 | 300.1 | Mixed anxiety and depressive disorder |
| ANX | F41.3 | ICD10 | 300.1 | Other mixed anxiety disorders |
| ANX | F41.8 | ICD10 | 300.1 | Other specified anxiety disorders |
| ANX | F41.9 | ICD10 | 300.1 | Anxiety disorder, unspecified |
| ANX | F94.0 | ICD10 | 313 | Selective mutism |

Supplemental Table 3. Temporal findings comparing phecodes in the EHR of those with isolated depression to those with isolated anxiety, considering the full available EHR, EHR truncated to before anxiety/depression diagnosis, and EHR limited to after anxiety/depression diagnosis.

compare_phewas_results.xlsx

N=8447. Patients with comorbid depression and anxiety were excluded. Phecodes were restricted to those that were significant in the Phe^2^WAS comparing patients with isolated depression vs. those with isolated anxiety in the analysis based on all available EHR data. OR_all is the OR from the full EHR Phe^2^WAS, where OR<1 indicates that the phecode is more common in isolated anxiety than isolated depression and OR>1 indicates that the phecode is more common with isolated depression than isolated anxiety. OR_before and P_before are the OR and p-value from the Phe^2^WAS limited to phecodes occurring before first anxiety/depression diagnosis. OR_after and P_after are the OR and p-value from the Phe^2^WAS limited to phecodes occurring after the first anxiety/depression diagnosis. OR_all may be compared to OR_before or OR_after to yield the relative strength of association between a medical diagnosis occurring either before or after a first diagnosis of depression/anxiety. For example, for phecode 278.11 morbid obesity, OR_all=2.10, but OR_after=2.43 suggesting that the association between morbid obesity with isolated depression over isolated anxiety was more pronounced for morbid obesity diagnoses made after first depression or anxiety diagnosis. P_diff_before_v_after is the p-value comparing the estimated effect of our phenotype (isolated depression vs isolated anxiety) with the phecode in the Phe^2^WAS truncated before diagnosis to the Phe^2^WAS truncated after diagnosis. P_diff_before_v_after compares the strength of association between a medical diagnosis and isolated depression or isolated anxiety in the EHR before first depression/anxiety diagnosis and the EHR after to first depression/anxiety diagnosis.

## Notes

### Competing Interest Statement

The authors have declared no competing interest.

### Author Declarations

This study was approved by the Mayo Clinic Institutional Review Board and the Mayo Clinic Biobank Access Committee.

